# Effects of tcVNS and Multimodal Stimulation on Heart Rate Variability and Stress Regulation

**DOI:** 10.1101/2025.06.22.25329895

**Authors:** Sungmin Ha, Seoyoung Choi, Yunjeong Jung, Sumin Lee, Woong Park

**Affiliations:** Graduate Program in Cognitive Science, Yonsei University, Seoul, Republic of Korea; Department of Psychology, Yonsei University, Seoul, Republic of Korea; Department of Physical Education, Yonsei University, Seoul, Republic of Korea; Department of Business Administration, Yonsei University, Seoul, Republic of Korea; Alus Healthcare Inc., Seoul, Republic of Korea

## Abstract

**Background:** Transcutaneous cervical vagus nerve stimulation (tcVNS) is a non-invasive neuromodulation that can influence autonomic function and emotional regulation by activating vagal pathways. Similarly, slow breathing exercises and aromatherapy are likewise known to enhance parasympathetic activity and reduce stress. However, the combined effects of multimodal sensory neuromodulation (e.g., combining electrical, mechanical respiratory, and olfactory stimuli) on autonomic nervous system activity, emotional stress, and prefrontal cortical activation remain underexplored. Further research is needed to clarify its potential benefits for stress regulation.

**Objective:** We investigated the effects of tcVNS, breathing training, and aroma inhalation—individually and combined—on heart rate variability (HRV), self-reported emotional stress, and prefrontal cortex (PFC) activation measured by functional near-infrared spectroscopy (fNIRS).

**Methods:** Twenty-three healthy adult participants underwent four conditions in a within-subject design: (1) Resting (baseline control, 5 minutes), (2) tcVNS stimulation (a custom-built transcutaneous cervical vagus nerve stimulator developed in-house at 25 Hz, 15 minutes) alone, (3) breathing + aroma (deep breathing exercise with calming aroma inhalation, 5 minutes) without tcVNS, and (4) combined tcVNS + breathing + aroma (15 minutes). Each condition lasted the specified duration, during which HRV indices (SDNN, RMSSD, pNN50, Poincaré SD1 and SD2, LF/HF ratio, approximate entropy (ApEn), and short-term fractal α_1_) were measured from PPG recordings, and changes in PFC oxyhemoglobin concentration were monitored with NIRSIT Quest (OBELAB, Seoul, South Korea) fNIRS system. Participants rated their emotional stress (state anxiety) after each condition. Statistical analyses used repeated-measures ANOVA and paired t-tests (with false discovery rate correction) to compare conditions.

**Results:** Both tcVNS and breathing+aroma conditions produced significant increases in HRV compared to resting baseline, and the combined tcVNS+breathing+aroma yielded the largest HRV enhancements. For example, SDNN (heart period variability) increased from 76.5±15.2 ms at rest to 109.5±18.7 ms with tcVNS (p=0.041, *q*=0.049) and to 121.1±20.3 ms with breathing+aroma (p<0.001, *q*=0.000), while the combined stimulation raised SDNN to 157.6±25.4 ms (p<0.001, *q*=0.001). Similar patterns were found in RMSSD, pNN50, SD1, and SD2, indicating augmented vagal tone. The combined condition significantly exceeded tcVNS-alone for these HRV metrics (all *q*<0.01). ApEn of HRV increased with tcVNS (from 1.05±0.10 to 1.23±0.14, p=0.006) and combined stimulation (*q*=0.052 vs. baseline), suggesting greater complexity under vagal stimulation, whereas breathing+aroma alone did not change ApEn. Self-reported emotional stress significantly decreased after all interventions, with the largest reduction in the combined tcVNS+breathing+aroma condition. Participants reported lower anxiety and worry, both remaining significant after FDR correction (q = .042). Additional reductions were observed in tension, restlessness, low mood, and stress reactivity (uncorrected p < .05), indicating a broad alleviation of negative affect consistent with the physiological findings. fNIRS revealed that breathing+aroma elicited increased PFC oxygenation bilaterally (mean Δoxy-Hb +0.11 µM, *p*<0.01 vs. rest), and combined stimulation produced an even larger PFC oxy-Hb increase (+0.19 µM, *p*<0.001). tcVNS alone caused a mild PFC activation (+0.05 µM, n.s.). PFC activation in the combined condition was significantly greater than in tcVNS alone and in breathing+aroma alone (*p*<0.05).

**Conclusions:** Multimodal stimulation combining tcVNS with breathing and aroma yielded synergistic benefits, evidenced by robust increases in vagally mediated HRV, lower emotional stress, and heightened PFC activity. These findings suggest that engaging both peripheral and central pathways through combined neuromodulatory and behavioral interventions can potentiate autonomic regulation and stress reduction beyond what single modalities achieve. This novel approach highlights a promising, non-pharmacological strategy for enhancing stress resilience and emotional well-being via coordinated autonomic and prefrontal modulation.

## Introduction

Chronic stress is widely recognized to disrupt the balance of the autonomic nervous system (ANS), typically manifesting as excessive sympathetic activity coupled with blunted parasympathetic tone (Won & Kim, 2016). One hallmark of this imbalance is a reduction in heart rate variability (HRV) – the beat-to-beat fluctuation in cardiac interbeat intervals – reflecting diminished vagal (parasympathetic) modulation of the heart. Under acute stress, high-frequency (HF) HRV components (associated with respiratory sinus arrhythmia and vagal activity) decrease while low-frequency (LF) components (linked to baroreflex-mediated sympathetic modulation) increase, indicating a shift toward sympathetic dominance (Kim et al., 2018). Conversely, greater HRV (for example, higher values of time-domain indices such as SDNN or RMSSD) is associated with better stress resilience and more adaptive emotional regulation capacity (Forte & Casagrande, 2025). The neurovisceral integration model posits that prefrontal cortical networks exert an inhibitory influence on subcortical autonomic centers, thereby maintaining flexible vagal control of the heart during stress and facilitating emotion regulation. Consistent with this model, neuroimaging studies have found that individuals with higher HRV exhibit increased activation of the ventromedial prefrontal cortex (vmPFC) during stress appraisal (Thayer et al., 2012). Thus, HRV serves not only as a peripheral index of cardiac autonomic balance but also as a proxy of central–peripheral integration in stress responses.

Transcutaneous cervical vagus nerve stimulation (tcVNS) has emerged as a promising non-invasive neuromodulation technique to enhance vagal activity and mitigate stress-related autonomic dysregulation. tcVNS delivers mild electrical stimulation to the cervical branch of the vagus nerve through surface electrodes on the neck, activating vagal afferent fibers that project to the nucleus tractus solitarius (NTS) in the brainstem (Shao et al., 2023). This intervention is known to acutely increase cardiovagal output and elevate HRV in both clinical and healthy populations. For example, in healthy young adults, transcutaneous auricular VNS (an anatomically analogous approach targeting the auricular branch) significantly increased vagally mediated HRV indices (e.g., RMSSD, pNN50, SDNN, HF power) and decreased the LF/HF ratio compared to sham stimulation, indicative of a shift toward parasympathetic predominance (Forte et al., 2022). Similarly, in a recent trial on patients with post-traumatic stress disorder (PTSD), active tcVNS (versus sham) attenuated stress-evoked sympathetic responses, including reducing the surge in heart rate and peripheral vascular resistance during traumatic stress exposure, effectively promoting an acute parasympathetic shift (Gurel et al., 2020). These findings underscore the mechanistic importance of tcVNS in suppressing sympathetic overdrive and enhancing vagal tone under stress, supporting its emerging therapeutic value as a stress-regulating intervention. Beyond its peripheral autonomic effects, vagus nerve stimulation also engages central pathways involved in affective regulation. Afferent vagal activation can influence limbic and cortical regions subserving emotion, and notably, transcutaneous auricular VNS has been shown to increase oxygenated hemoglobin levels in the prefrontal cortex (PFC) and to strengthen prefrontal functional connectivity during stimulation (Zhu et al., 2024;). This PFC engagement via vagal pathways aligns with reports of improved emotional regulation and executive control in some VNS studies. Taken together, tcVNS appears capable of concurrently modulating peripheral autonomic outflow and recruiting frontal neural circuits that support adaptive stress responses, making it a compelling tool for autonomic and affective regulation.

Slow, controlled breathing is another powerful modulator of the ANS, operating through baroreflex and cardiopulmonary coupling mechanisms. Breathing at approximately six breaths per minute (∼0.1 Hz) maximizes respiratory sinus arrhythmia and has been shown to increase vagal cardiac control while reducing sympathetic activity (Russo et al., 2017; Lehrer et al., 2020). Even a single session of deep, slow breathing can significantly elevate high-frequency HRV and decrease state anxiety levels. For instance, Magnon et al. (2021) demonstrated that a 5-minute session of paced abdominal breathing (with prolonged exhalation at ∼6 breaths per minute) tended to increase vagally mediated HRV and significantly reduced state anxiety, particularly in older adults. These findings suggest that even brief sessions of slow, paced breathing can enhance vagally mediated autonomic function and reduce anxiety particularly in older adults although the underlying mechanisms, such as baroreflex involvement, require further investigation.

Beyond these bottom-up physiological effects, breathing exercises also provide a top-down cognitive anchor that can alleviate emotional distress (Balban et al., 2023). Breathing techniques are frequently employed in mindfulness and relaxation practices to induce calm and reduce perceived stress, and regular practice of slow diaphragmatic breathing has been associated with lower basal cortisol levels and improvements in mood and cognitive function (Ma et al., 2017). The act of focusing on one’s breath engages frontal brain regions involved in attention and interoceptive awareness. Consistent with this, functional neuroimaging studies on meditation and attentional tasks have reported increased activation and cerebral blood flow in the prefrontal cortex, including the ventrolateral PFC. Although direct neuroimaging evidence on isolated breathing is limited, these findings suggest that focused breathing may recruit prefrontal areas involved in top-down attentional control (Braboszcz et al., 2010). Thus, paced breathing provides both a direct autonomic calming effect and a cognitive means of promoting emotion regulation.

Aromatherapy with calming essential oils represents a complementary sensory approach to stress reduction that can be integrated with physiological interventions. Inhalation of certain fragrances, such as bergamot, lavender, and chamomile, has been associated with anxiolytic effects and modest increases in parasympathetic activity. Olfactory stimuli have direct access to limbic structures like the amygdala and hippocampus, allowing scents to rapidly influence emotional states (Chien et al., 2012; Watanabe et al., 2015; Agarwal et al., 2022). Bergamot (Citrus bergamia), in particular, has been studied for its potential to reduce anxiety and impact autonomic parameters. In a randomized crossover study, Watanabe et al. (2015) found that healthy female participants who inhaled bergamot essential oil experienced significant increases in high-frequency (HF) heart rate variability (HRV) and decreases in salivary cortisol levels, indicating enhanced vagal modulation and reduced stress. These physiological changes coincided with improved mood states and lower anxiety levels. Other reports similarly note that pleasant aromas can facilitate parasympathetic recovery following stress and increase overall HRV during relaxation (Chung et al., 2024; Kim, Lee, & Song, 2024).

The relaxation effects of aromatherapy are thought to operate via both psychological pathways (e.g., positive affect and reduced perceived stress) and neurophysiological pathways (olfactory activation of limbic calming circuits). While the autonomic impact of aroma inhalation alone is often smaller than that produced by techniques like slow breathing, the addition of a soothing scent can augment the overall relaxation response. For example, in a study by Chang and Shen (2011), participants who inhaled bergamot essential oil demonstrated greater parasympathetic recovery following stress, as indicated by increased high-frequency HRV and reduced heart rate. These changes suggest enhanced vagal modulation and improved stress resilience. Thus, incorporating aromatherapy creates an enriched, multisensory environment that facilitates the down-regulation of stress and sympathetic arousal.

Given that tcVNS, slow breathing, and aromatherapy each can independently promote parasympathetic activity and stress relief, a logical question arises as to whether combining these modalities yields additive or synergistic benefits. Multimodal interventions targeting both peripheral autonomic pathways and central sensory/cognitive pathways could potentially reinforce each other’s effects on the stress response. There is some precedent for pairing vagus nerve stimulation with respiratory maneuvers: for example, research on respiratory-gated auricular VNS (RAVANS), which times VNS pulses to the exhalation phase of breathing, has suggested enhanced baroreflex engagement and vagal outflow, although findings have been mixed (Sclocco et al., 2020; Garcia et al., 2022; Szulczewski et al., 2023). Early studies reported that concurrent slow breathing and transcutaneous VNS produced greater vagal cardiac control than breathing alone, but later controlled experiments found that transcutaneous auricular VNS did not significantly augment HRV beyond the effects of slow breathing itself (Paccione et al., 2022). These inconsistent outcomes underscore the need to clarify how combined vagal stimulation and respiratory interventions interact. Notably, prior work on RAVANS focused on auricular VNS and did not incorporate additional calming modalities like olfactory stimuli, nor did it measure central indicators of arousal regulation such as cortical activity. Thus, multimodal sensory neuromodulation (e.g., combining electrical, mechanical respiratory, and olfactory stimuli) remains an emerging area of study, and its potential benefits for stress regulation have yet to be fully elucidated.

In light of this gap, the present study systematically evaluated the acute effects of tcVNS, slow breathing, and aroma inhalation—alone and in combination—on autonomic and emotional outcomes. Multimodal sensory neuromodulation (e.g. combining electrical, respiratory, and olfactory stimuli) remains an emerging area of study. We hypothesized that integrating tcVNS with paced breathing and aromatherapy would engage multiple pathways of calming – afferent vagal signaling, baroreflex enhancement, and limbic soothing – leading to greater overall autonomic and emotional regulation than any single modality alone. To our knowledge, this is the first experimental evaluation of cervical VNS combined with behavioral and sensory interventions for acute stress modulation. We employed a within-subject experimental design in which 23 healthy adults underwent four conditions: a resting baseline, tcVNS stimulation alone, breathing + aroma (deep breathing with calming aroma inhalation) alone, and combined tcVNS + breathing + aroma. During each condition, we recorded a comprehensive panel of HRV metrics—including time-domain, frequency-domain, and non-linear indices of vagal activity—using a PPG sensor placed on the index finger of the non-dominant hand. We also obtained self-reported emotional stress ratings and monitored prefrontal cortex hemodynamic activity using functional near-infrared spectroscopy (fNIRS). We hypothesized that each active intervention would acutely increase vagal-mediated HRV and reduce subjective stress compared to baseline, and that the combined tcVNS+breathing+aroma would engage convergent autonomic and neurocognitive mechanisms to produce the greatest enhancement of parasympathetic activity and the largest reduction in stress, surpassing the effect of either modality alone. Furthermore, given the central role of the PFC in autonomic regulation, we expected PFC oxygenation to increase during the active conditions, with the combined stimulation yielding the most robust PFC activation in line with its hypothesized synergistic effect. By elucidating the physiological and neural impacts of this multimodal approach, our study aims to inform the development of integrated mind-body interventions for stress and anxiety, offering a novel non-pharmacological strategy to enhance stress resilience and emotional well-being via coordinated autonomic and prefrontal modulation.

## Methods

### 1. Participants

Twenty-three healthy adults (13 female, 10 male; age 21.0 ± 2.1 years), all undergraduate students from Yonsei University and right-handed, participated in this study. All participants were free of cardiovascular, respiratory, neurological, or psychiatric disorders and were not on medications known to affect heart rate or autonomic function. Written informed consent was obtained from each participant, and the protocol was approved by the Institutional Review Board of Yonsei University (*7001988-202412-HR-2369-02*)

### 2. Experimental Design and Procedures

We employed a within-subject, repeated-measures experimental design in which each participant underwent four conditions: Resting baseline, tcVNS alone, Breathing+Aroma, and Combined tcVNS + Breathing + Aroma. The overall procedure, including the order of conditions and surveys, is illustrated in Figure 1 (right). The order of conditions was fixed across participants to control for residual scent effects and ensure experimental consistency, with aroma-containing conditions placed later in the sequence to avoid unintended olfactory carryover. All sessions took place in a quiet, dimly lit room. Prior to the start of the experimental session, all physiological sensors were attached, including the PPG device for HRV measurement, the fNIRS optodes for cortical monitoring, and the tcVNS electrodes for stimulation.

**Figure 1.**
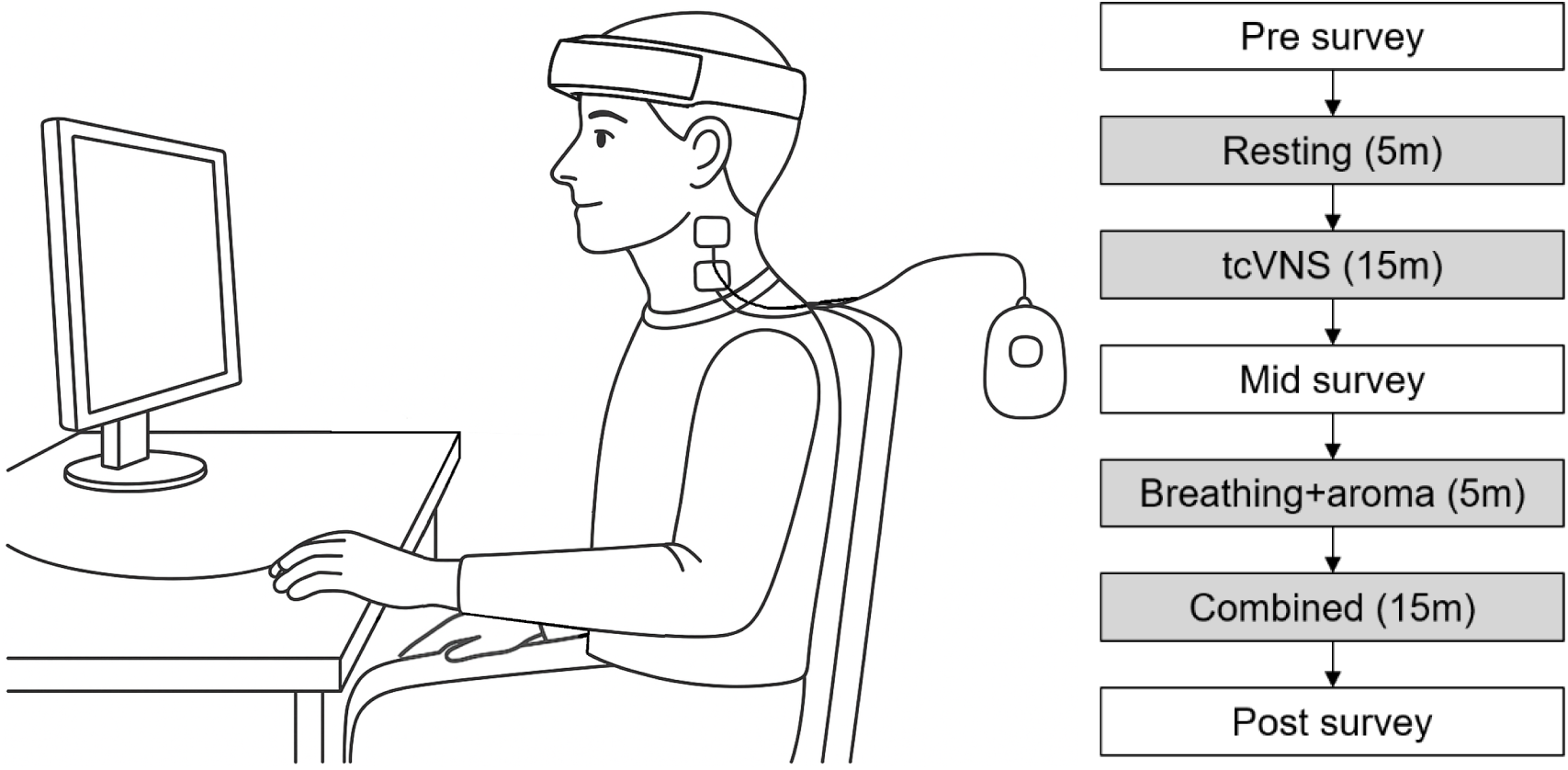
Electrode placement for tcVNS (left) and the experimental procedure timeline including all stimulation and survey phases (right).

During the Resting baseline condition, participants remained seated in a comfortable chair for five minutes without any active intervention. They were instructed to maintain a relaxed posture, breathe naturally, and visually fixate on a white cross presented at the center of a black screen. This condition served to establish a physiological and neural baseline for heart rate variability (HRV), perceived stress, and cortical hemodynamic activity under passive resting conditions.

In the tcVNS condition, participants received transcutaneous cervical vagus nerve stimulation while sitting at rest (15 minutes). The tcVNS was delivered using a custom-designed, lab-developed portable stimulator (see Appendix A for design details), with surface electrodes placed on the skin over the left cervical vagus nerve (side of the neck). Electrodes were positioned between the sternocleidomastoid and trachea at the cricoid cartilage level, following standard anatomical landmarks, as shown in the left panel of Figure 1. Stimulation parameters were set to 25 Hz, 400 µs pulse width, and a fixed peak amplitude of ±3.5 V. The tcVNS device was kept on continuously for the 15-minute period. During tcVNS, participants simply rested while fixating on a central cross. A trained researcher monitored for any signs of discomfort or adverse effects. No significant side effects were reported aside from the expected tingling at the stimulation site.

In the Breathing + Aroma condition, participants practiced guided slow breathing while being exposed to a calming aroma, but without any electrical stimulation (5 minutes). They were instructed in a diaphragmatic breathing technique: inhale deeply through the nose for ∼4 seconds, hold the breath for ∼2 seconds, then exhale slowly for ∼4 seconds (target breathing rate ∼6 breaths/min). A visual breathing guide was provided to help maintain a consistent pace. Participants briefly practiced the breathing technique twice before the experimental session to ensure consistency. The bergamot-scented perfume was presented in a manner designed to minimize participant awareness of its delivery: a mild concentration was sprayed onto a piece of paper placed approximately 1 meter behind the participant, outside their direct visual field. This ensured that olfactory stimulation coincided with inhalation without inducing expectancy effects. We used a bergamot perfume for its well-documented anxiolytic properties; the choice of bergamot was motivated by prior evidence of its autonomic and mood benefits (Watanabe et al., 2015). This condition was intended to activate the parasympathetic nervous system through respiratory sinus arrhythmia and induce relaxation through olfactory pathways.

In the Combined tcVNS + Breathing + roma condition (15 minutes), all three interventions were applied simultaneously. Participants received tcVNS (same device settings as the tcVNS-alone condition) while performing the guided slow diaphragmatic breathing and inhaling the bergamot aroma. The setup was essentially a merger of the above two conditions: the tcVNS stimulation commenced, and participants followed the same breathing pacing cues as in the Breathing + Aroma condition. The rationale was to engage vagal afferents (via electrical stimulation) in concert with the natural vagal enhancement from breathing, under a soothing sensory context. Research staff closely observed the participants for any signs of discomfort due to the combined stimuli. All participants were able to complete the combined session without difficulties.

Throughout all conditions, participants were seated in the same position and instructed to minimize movement and avoid speaking to prevent motion artifacts in the physiological recordings. Before each experimental measurement, a calibration period was provided to ensure that the fNIRS and PPG data were being properly recorded and that heart rate and breathing were stable. At the end of each 15-minute condition, we obtained self-report measurements before transitioning to the next condition (or ending the session). The experimenter remained in an adjacent area during the conditions, out of the participant’s direct view, to reduce any observer effect, but available immediately if the participant indicated any need to stop.

### 3. Measures and Data Analysis

#### Heart Rate Variability (HRV)

Heart activity was monitored via photoplethysmography (PPG) to derive heart rate and heart rate variability (HRV) metrics, which serve as noninvasive indices of autonomic nervous system regulation and psychophysiological flexibility. A PPG sensor was affixed to the index finger of the participant’s non-dominant hand. The PPG signal was sampled at 100 Hz and synchronized with the fNIRS system. Inter-beat intervals (IBIs) were extracted from the pulse waveform, and R-R intervals (RRi) were detected using an automated peak-detection algorithm followed by manual correction to ensure fidelity. From the RRi time series, we calculated several HRV indices with distinct physiological interpretations:

● SDNN (ms), the standard deviation of NN intervals, reflects overall HRV and is sensitive to both sympathetic and parasympathetic influences; higher SDNN is typically associated with greater cardiovascular adaptability and lower stress burden.
● RMSSD (ms), the root-mean-square of successive RR differences, captures short-term vagal modulation; elevated RMSSD is considered a robust marker of parasympathetic (vagal) activity and emotional regulation capacity.
● pNN50 (%), the percentage of adjacent RR intervals differing by more than 50 ms, also indexes vagal tone, with higher values reflecting greater beat-to-beat variability and stress resilience.

Nonlinear geometric indices derived from Poincaré plots were also analyzed:

● SD1 (ms) reflects instantaneous short-term variability, closely related to RMSSD and vagal input.
● SD2 (ms) captures long-term variability and is influenced by both branches of the autonomic nervous system.

Frequency-domain features were obtained using power spectral density estimation (Welch’s method) applied to interpolated RRi signals resampled at 4 Hz. In particular, we examined the LF/HF ratio, defined as the ratio of low-frequency power (0.04–0.15 Hz) to high-frequency power (0.15–0.4 Hz). Although this index is sometimes interpreted as a marker of sympathovagal balance, its interpretation requires caution under fixed breathing rates (∼0.1 Hz), as used in our protocol.

To assess signal complexity and dynamical structure, we also computed:

● Approximate Entropy (ApEn), which quantifies the unpredictability or complexity of the RRi series; higher ApEn values suggest more adaptable and flexible heart rate patterns, while lower values may indicate rigid or overly structured autonomic output.
● DFA-α₁, the short-term scaling exponent from Detrended Fluctuation Analysis, captures fractal-like correlations in heart rhythm fluctuations over short timescales. Values approaching 1.0 are considered optimal and reflect healthy intrinsic autonomic regulation.

Together, these HRV indices provide a multidimensional characterization of cardiac autonomic control, capturing both vagal responsiveness and system complexity, which are critical for physiological homeostasis and psychological resilience. All HRV analyses were conducted using the NeuroKit2 Python library (v0.2.4). R-peaks were extracted from cleaned PPG signals, and RR intervals were computed accordingly. Basic artifact correction and peak validation were applied during preprocessing.

#### Emotional Stress Assessment

Subjective emotional stress was assessed using the Korean Emotional Stress Scale developed by Jeon et al. (2020), which measures six domains of negative emotional stress: state anger, trait anger, state anxiety, trait anxiety, state depression, and trait depression. These represent both situational (state) and dispositional (trait) components of anger, anxiety, and depression. The validated instrument consists of six subscales, each comprising seven items rated on a 5-point Likert scale (1 = not at all, 5 = very much). Participants were instructed to rate how they felt “at this moment” immediately after completing each experimental condition for state subscales, while trait subscales reflect more general and enduring emotional tendencies. Subscale scores were calculated by summing responses within each domain. A composite emotional stress index was also computed by averaging the six subscale scores, with higher scores indicating greater emotional stress.

In addition to analyzing aggregated subscale scores, we examined item-level changes to capture more nuanced emotional dynamics. Participants completed all 23 items before (pre) and after (post) each condition allowing for within-subject comparisons at the level of individual affective descriptors (e.g., “irritable,” “anxious,” “hopeless”). This enabled us to detect specific emotional shifts that may not be reflected in broader subscale averages, particularly in response to targeted interventions. We focused our analysis on differences in post-condition state stress ratings relative to baseline, and applied paired-sample t-tests with False Discovery Rate (FDR) correction to evaluate statistical significance at the item level. The scale has demonstrated high internal consistency (Cronbach’s α = .86–.93 for state subscales) and good convergent validity with other standardized measures such as the STAI, STAXI, and CES-D (Jeon et al., 2020).

#### Prefrontal Cortex Activation (fNIRS)

Cerebral hemodynamic activity was recorded with functional near-infrared spectroscopy. We used the NIRSIT Quest (OBELAB, Seoul, South Korea) fNIRS system, a high-density, wireless multi channel device. The NIRSIT Quest employs near-infrared light at two wavelengths (approximately 780 nm and 850 nm) and contains an array of sources and detectors configured to measure frontal cortical regions. This allowed calculation of concentration changes in oxygenated hemoglobin (oxy-Hb) and deoxygenated hemoglobin (deoxy-Hb) using the modified Beer-Lambert law, with differential pathlength factors applied for adult forehead tissue. We were primarily interested in oxy-Hb changes, as increases in oxy-Hb reflect elevated regional cerebral blood flow and neural activation.

Before each session, the fNIRS optode cap was placed and calibrated to ensure good signal quality (low optical noise and sufficient gain on all channels). The device was placed over the participant’s forehead, covering the bilateral prefrontal cortex area (see Figure 3). In its standard configuration, NIRSIT Quest yields up to 48 measurement channels (source-detector pairs) at a sampling rate of ∼8.13 Hz, with source-detector separations optimized for cortical sensitivity. Before starting the experiment, the fNIRS cap was fitted snugly and checked for proper optode-skin contact and signal quality. The fNIRS data acquisition software recorded the raw light intensity signals for each channel continuously across the entire session. Event markers were logged in real time to indicate the onset and offset of each condition, enabling precise segmentation for later analysis.

**Figure 3.**
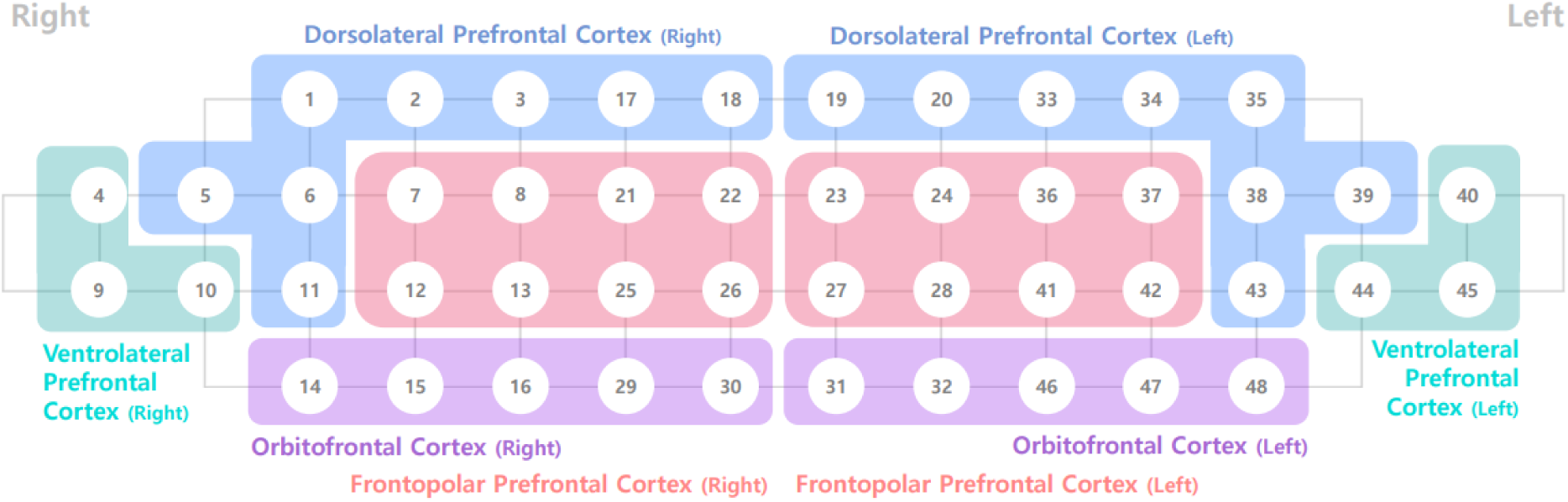
fNIRS channel layout and optode configuration for prefrontal cortex measurement using NIRSIT Quest.

### 4. Statistical Analysis

We analyzed the data using repeated-measures statistics with the within-subject factor Condition (four levels: Resting, tcVNS alone, Resting+Breathing+Aroma, tcVNS+Breathing+Aroma). For each dependent variable (each HRV index, the stress rating, and PFC oxy-Hb), we first conducted a one-way repeated-measures ANOVA to test for any overall effect of Condition. When Mauchly’s test indicated non-sphericity, Greenhouse-Geisser corrections were applied to *p*-values. A significant ANOVA (α = 0.05) was followed by planned pairwise paired t-tests between specific conditions to identify differences. In particular, we had a priori interest in comparisons of each active condition to baseline resting condition and the synergy-related comparisons. We applied a False Discovery Rate (FDR) correction to adjust for multiple comparisons within each family of tests (Benjamini-Hochberg procedure), controlling the FDR at 0.05. All statistical analyses were conducted using Python’s SciPy library. Significance was defined as *p* < 0.05 (two-tailed) after correction. Data are presented as mean ± SD in text.

To evaluate regional hemodynamic differences between conditions, statistical analyses were performed on the fNIRS-derived changes in oxygenated (HbO) and deoxygenated hemoglobin (HbR) for each channel. Given the relatively small sample size and the non-normal distribution of several channel-wise data (as assessed by Shapiro-Wilk tests), non-parametric Wilcoxon signed-rank tests were conducted to compare the experimental conditions (e.g., tcVNS condition against the resting baseline) for each channel separately. This test evaluates whether the median paired differences between conditions deviate significantly from zero without assuming normality. Analyses were conducted independently for HbO and HbR signals. All p-values reported are uncorrected for multiple comparisons unless otherwise noted. Channels showing p < .05 were identified as preliminarily significant and interpreted in light of their spatial location and consistency with expected prefrontal activation patterns.

## Results

### 1. Heart Rate Variability

Heart rate variability indices were analyzed across four experimental conditions: Resting, tcVNS, Breathing + Aroma, and Combined (tcVNS + Breathing + Aroma). A series of pairwise comparisons were conducted, and all reported p-values were corrected using the Benjamini-Hochberg False Discovery Rate (FDR) method. Significant effects were observed across multiple HRV domains, indicating distinct autonomic modulations induced by each intervention.

#### SDNN (Standard Deviation of NN intervals)

Compared to Resting (M = 76.52 ms), SDNN significantly increased in the tcVNS condition (M = 109.49 ms; t(20) = –2.18, p = .041, q = .049), and even more so in the Breathing + Aroma (M = 121.07 ms; t(20) = –4.66, q = .000) and Combined condition (M = 157.56 ms; t(20) = –4.22, q = .001) as shown in Figure 4. Combined was also significantly greater than tcVNS (t(20) = –4.65, q = .000) and Breathing + Aroma (t(20) = –2.53, q = .030).

**Figure 4.**
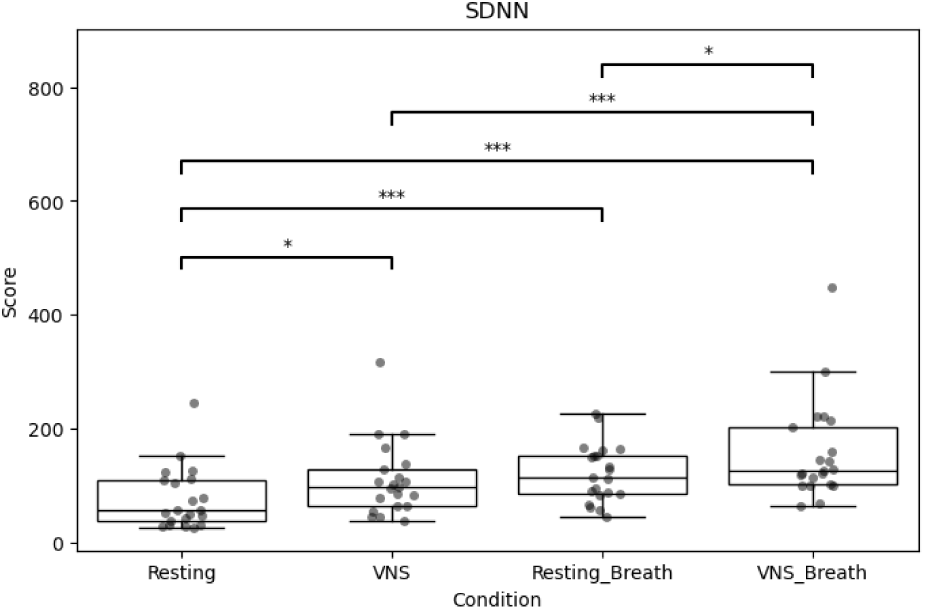
Group differences in SDNN across experimental conditions.

#### RMSSD (Root Mean Square of Successive Differences)

RMSSD increased from 87.09 ms at baseline to 131.72 ms in tcVNS (t(20) = –2.16, q = .051), 139.40 ms in Breathing + Aroma (t(20) = –3.91, q = .002), and 195.45 ms in the Combined condition (t(20) = –4.14, q = .002), as shown in Figure 5. Combined was significantly higher than both tcVNS (t(20) = –4.25, q = .002) and Breathing + Aroma (t(20) = –2.98, q = .011).

**Figure 5.**
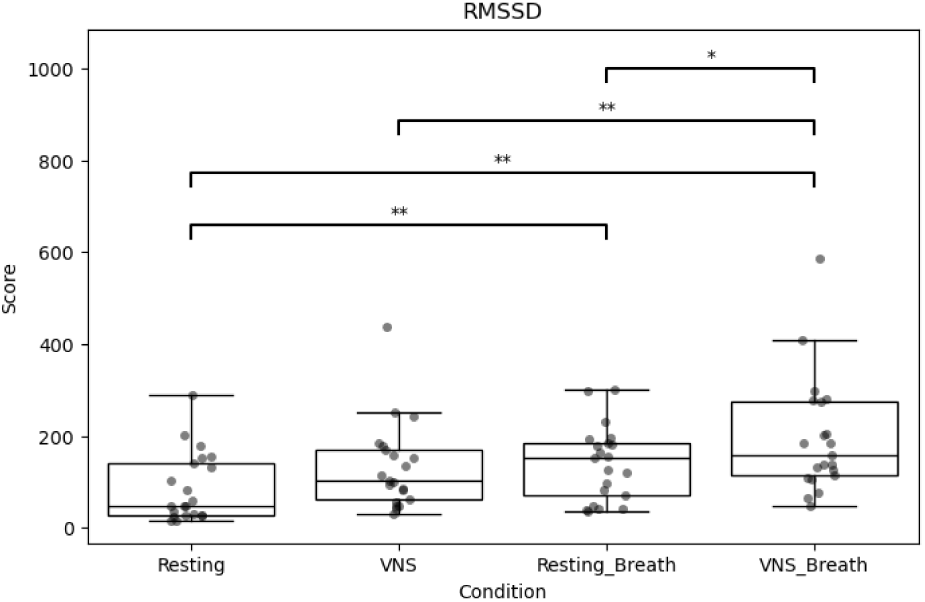
Group differences in RMSSD across experimental conditions.

#### pNN50 (percentage of adjacent RR intervals differing >50 ms)

pNN50 increased from 20.83% (Resting) to 30.12% in tcVNS (t(20) = –2.69, q = .021), 35.55% in Breathing + Aroma (t(20) = –4.05, q = .002), and 42.31% in the Combined condition (t(20) = –4.86, q = .001), as shown in Figure 6. Combined was significantly higher than tcVNS (t(20) = –3.69, q = .003).

**Figure 6.**
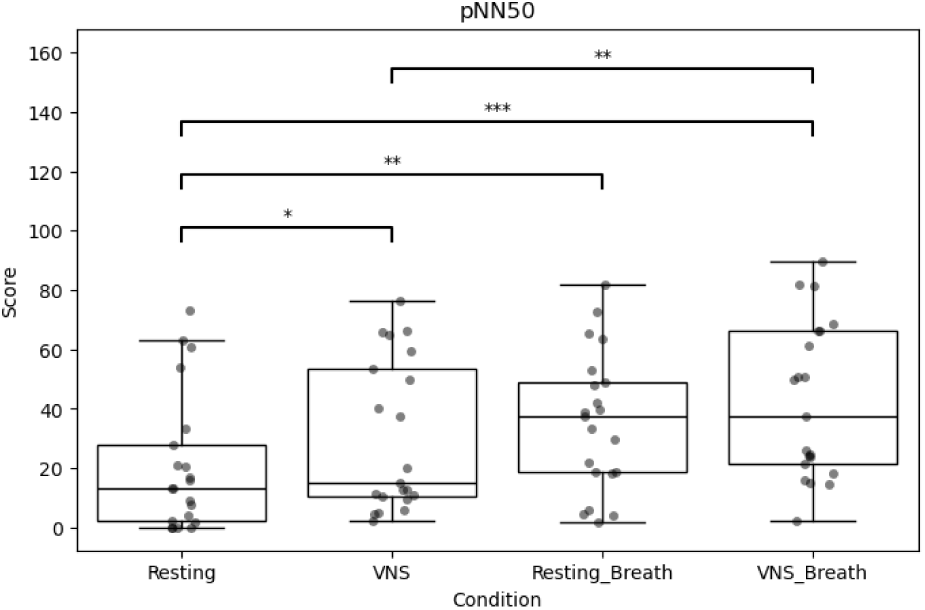
Group differences in pNN50 across experimental conditions.

#### Poincaré Plot Measures

SD1 rose from 61.67 ms at Resting to 93.21 ms in tcVNS (t(20) = –2.16, q = .052), 98.69 ms in Breathing + Aroma (t(20) = –3.91, q = .002), and 138.27 ms in Combined (t(20) = –4.14, q = .002), as shown in Figure 7. Combined was significantly higher than tcVNS (t(20) = –4.25, q = .002) and Breathing + Aroma (t(20) = –2.97, q = .011). SD2 increased significantly from Resting (87.52 ms) to tcVNS (122.22 ms; t(20) = –2.18, q = .050), Breathing + Aroma (138.07 ms; t(20) = –4.79, q = .000), and Combined (173.30 ms; t(20) = –4.19, q = .001), **as** shown in Figure 8. Combined was greater than both tcVNS (t(20) = –4.71, q = .000) and Breathing + Aroma (t(20) = –2.22, q = .050).

**Figure 7.**
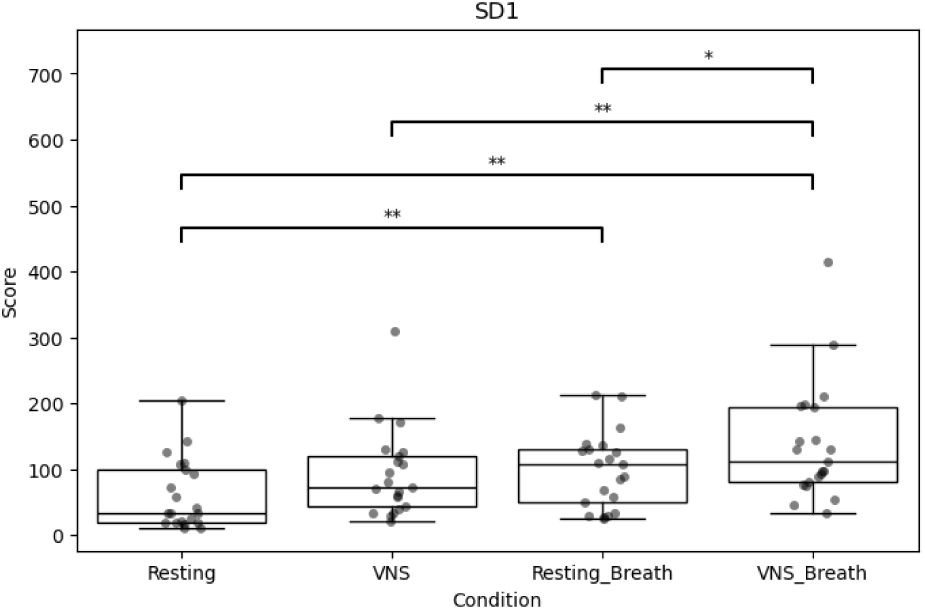
Group differences in SD1 across experimental conditions.

**Figure 8.**
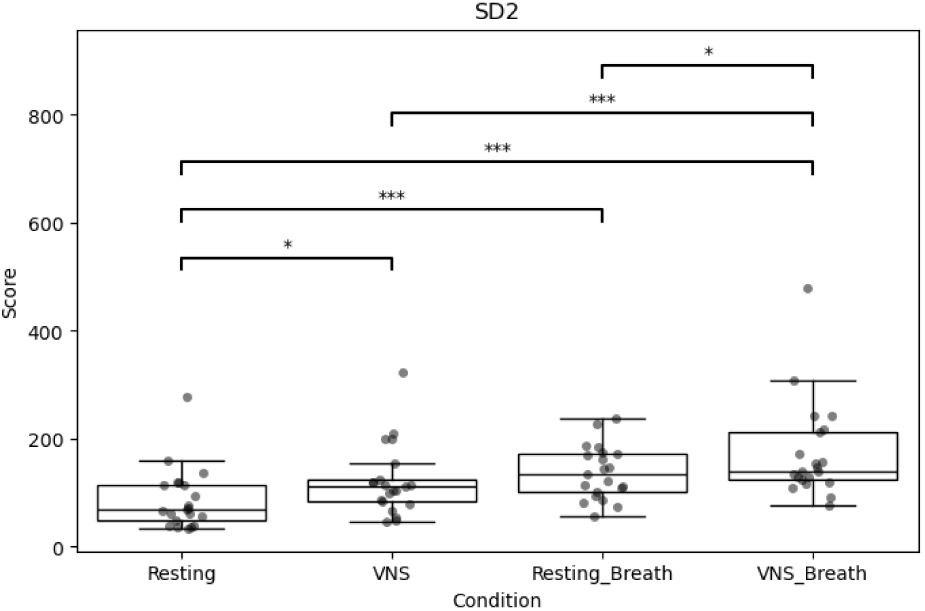
Group differences in SD2 across experimental conditions.

#### Approximate Entropy (ApEn)

ApEn increased from 1.05 (Resting) to 1.23 with tcVNS (t(20) = –3.06, q = .019), as shown in Figure 9, remained similar with Combined (1.23), and decreased slightly with Breathing + Aroma (1.04). tcVNS was significantly higher than Breathing + Aroma (t(20) = 2.87, q = .019), and Combined was also significantly higher than Breathing + Aroma (t(20) = –4.15, q = .003). The increase from Resting to Combined was marginal (t(20) = –2.26, q = .052).

**Figure 9.**
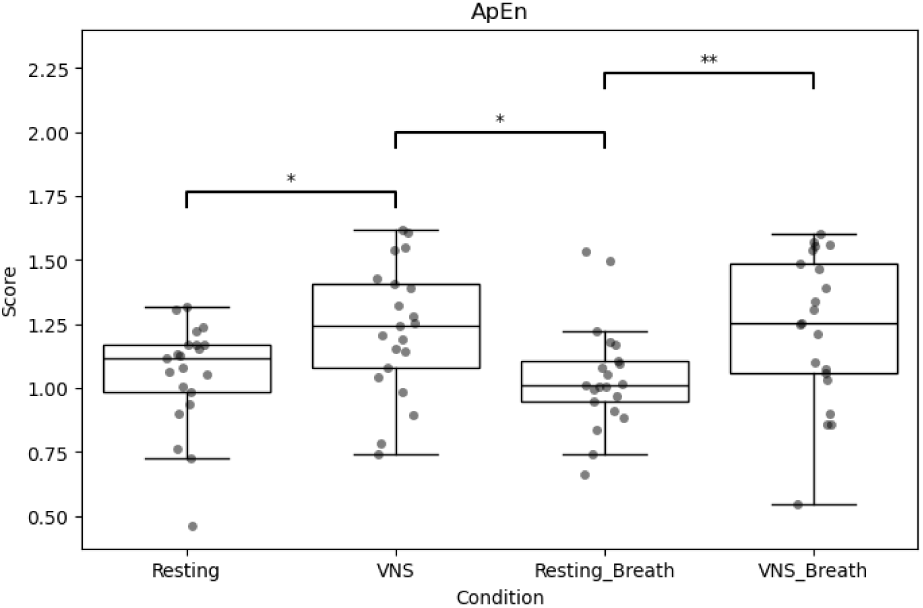
Group differences in ApEn across experimental conditions.

#### DFA-α₁ (Detrended Fluctuation Analysis, short-term scaling exponent)

DFA-α₁ increased significantly from Resting (0.76) to Breathing + Aroma (0.96; t(20) = –3.35, q = .019), indicating enhanced fractal correlation (see Figure 10). The increase in Combined (0.95) was not significantly different from Breathing + Aroma, and tcVNS alone (0.79) showed only a marginal increase compared to Breathing + Aroma (t(20) = –2.54, q = .058).

**Figure 10.**
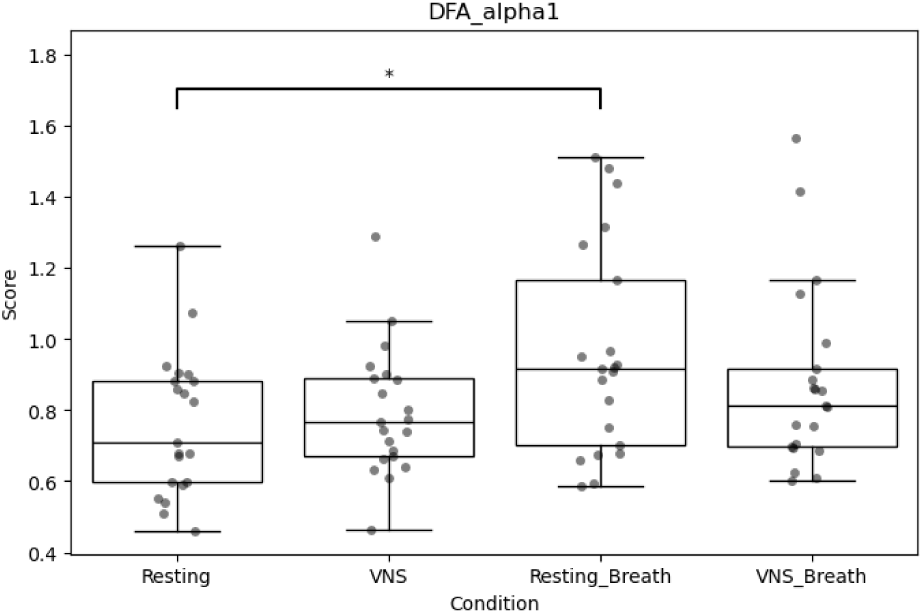
Group differences in DFA-α₁ across experimental conditions.

All three active interventions (tcVNS, Breathing + Aroma, and combined multimodal stimulation) led to measurable increases in heart rate variability relative to resting baseline, as reflected in time-domain (SDNN, RMSSD, pNN50), nonlinear (SD1, SD2), and complexity (ApEn) indices. Notably, tcVNS alone elicited significant autonomic changes, including increased SDNN, RMSSD, pNN50, and ApEn, indicating enhanced vagal activity and elevated complexity in cardiac regulation even in the absence of paced breathing or olfactory input. This supports the autonomic efficacy of tcVNS as a standalone intervention.

The combined multimodal condition, however, consistently yielded the largest effects (nearly doubling of SDNN and RMSSD, and the highest pNN50 and SD2 values across conditions) demonstrating a robust augmentation of both short and long term parasympathetic modulation, as detailed in Table 1. Approximate entropy also increased significantly with tcVNS and combined multimodal stimulation, but not with Breathing + Aroma, suggesting that electrical stimulation preserved or enhanced heart rate complexity, even during structured breathing. In contrast, the fractal scaling exponent (DFA-α₁) improved significantly only with Breathing + Aroma, indicating greater long-range order and self-similarity in autonomic control. Together, these findings underscore the independent effectiveness of tcVNS, the complementary contributions of respiratory-olfactory stimulation, and the synergistic benefit of combining them. The combined multimodal intervention thus achieved greater autonomic flexibility and vagal regulation than either component alone.

**Table 1.** Repeated-measures ANOVA and post-hoc comparisons of HRV indices across experimental conditions (FDR-corrected p-values).

| Measure | F (P value) | $\eta^2$ | Post-hoc FDR-corrected p | | | | | |
| --- | --- | --- | --- | --- | --- | --- | --- | --- |
|  |  |  | Resting vs. tcVNS | Resting vs. Breathing + aroma | Resting vs. Combined | tcVNS vs. Breathing + aroma | tcVNS vs. Combined | Breathing + aroma vs. Combined |
| SDNN | 11.254 (<.000) | 0.167 | 0.049* | 0.000* | 0.001* | 0.409 | 0.000* | 0.030* |
| RMSSD | 10.754 (<.000) | 0.144 | 0.051 | 0.002* | 0.002* | 0.682 | 0.002* | 0.011* |
| pNN50 | 10.404 (<.000) | 0.097 | 0.021* | 0.002* | 0.001* | 0.286 | 0.003* | 0.114 |
| LF | 0.917 (.438) | 0.020 | 0.466 | 0.466 | 0.734 | 0.803 | 0.875 | 0.815 |
| HF | 0.238 (.870) | 0.006 | 0.946 | 0.946 | 0.946 | 0.946 | 0.946 | 0.961 |
| LF/HF | 3.042 (.083) | 0.101 | 0.259 | 0.143 | 0.143 | 0.143 | 0.163 | 0.259 |
| SD1 | 10.725 (<.000) | 0.143 | 0.052 | 0.002* | 0.002* | 0.680 | 0.002* | 0.011* |
| SD2 | 11.076 (<.000) | 0.180 | 0.050* | 0.000* | 0.001* | 0.297 | 0.000* | 0.050* |
| ApEn | 5.395 (.002) | 0.134 | 0.019* | 0.946 | 0.052 | 0.019* | 0.946 | 0.003* |
| DFA-$\alpha_1$ | 4.348 (.008) | 0.100 | 0.505 | 0.019* | 0.168 | 0.058 | 0.237 | 0.237 |

### 2. Emotional Stress and Anxiety

To assess the emotional impact of the combined multimodal intervention (tcVNS + breathing + aroma), paired-sample *t*-tests were conducted on pre- and post-intervention scores for each item in the emotional stress survey. Several items exhibited statistically significant reductions in scores after the intervention (uncorrected *p* < .05), indicating an alleviation of subjective emotional distress, as summarized in Table 2. Participants reported significantly lower levels of anticipatory worry (*“I worry that misfortune is about to happen“*, M = 1.52 post vs. 2.17 pre, *t*(22) = 2.81, *p* = .010), tension (*“I feel tense“*, M = 1.91 vs. 2.57, *t*(22) = 2.13, *p* = .044), anxiety (*“I feel anxious“*, M = 1.57 vs. 2.43, *t*(22) = 3.79, *p* = .001), and restlessness (*“I feel restless“*, M = 1.57 vs. 2.04, *t*(22) = 2.55, *p* = .018). Additional improvements were observed in items reflecting depressive or demotivated states, including a reduced sense of life-weariness (*“I have no will to live“*, M = 1.26 vs. 1.61, *t*(22) = 2.15, *p* = .043), feelings of unhappiness (*“I feel unhappy“*, M = 1.35 vs. 1.74, *t*(22) = 2.40, *p* = .025), and a diminished sense of personal failure (*“I feel like a loser“*, M = 1.22 vs. 1.52, *t*(22) = 2.61, *p* = .016). Reactivity to minor stressors also showed reduction, with significantly lower scores on *“I flare up over trivial matters“* (M = 1.57 vs. 2.00, *t*(22) = 2.65, *p* = .015), *“I worry about trivial things“* (M = 1.96 vs. 3.00, *t*(22) = 3.43, *p* = .002), and *“I can’t shake off bad thoughts when they come“* (M = 1.96 vs. 2.61, *t*(22) = 2.34, *p* = .029). Lastly, participants reported lower negative mood on items such as *“I feel gloomy“* (M = 1.74 vs. 2.17, *t*(22) = 2.33, *p* = .030) and *“I feel impatient when I’m late for an appointment“* (M = 2.91 vs. 3.74, *t*(22) = 2.45, *p* = .022). Among all statistically significant items, the reductions in anxiety (“I feel anxious”, q = .042) and trivial worry (“I worry about trivial things”, q = .042) remained significant after correction for multiple comparisons using the Benjamini–Hochberg FDR procedure. This implies that while the combined multimodal intervention broadly reduced emotional reactivity and negative affect, its most robust and statistically reliable effect was the alleviation of momentary anxiety and generalized worry.

**Table 2.** Changes in self-reported emotional items before and after intervention: t-tests and FDR-corrected significance.

| Item | pre Mean | post Mean | t-value | p-value | FDR q-value |
| --- | --- | --- | --- | --- | --- |
| I worry that misfortune is about to happen. | 2.17 | 1.52 | 2.812 | 0.01* | 0.126 |
| I feel tense. | 2.57 | 1.91 | 2.135 | 0.044* | 0.154 |
| I feel anxious. | 2.43 | 1.57 | 3.792 | 0.001* | 0.042* |
| I feel restless. | 2.04 | 1.57 | 2.554 | 0.018* | 0.126 |
| I feel afraid. | 1.61 | 1.26 | 1.886 | 0.073 | 0.173 |
| My heart trembles. | 1.91 | 1.65 | 1.064 | 0.299 | 0.417 |
| I feel uneasy. | 1.78 | 1.61 | 0.848 | 0.406 | 0.487 |
| I feel hopeless. | 1.48 | 1.26 | 2.011 | 0.057 | 0.171 |
| I feel like a failure in life. | 1.52 | 1.35 | 1.447 | 0.162 | 0.279 |
| I have no will to live. | 1.61 | 1.26 | 2.152 | 0.043* | 0.154 |
| I feel unhappy. | 1.74 | 1.35 | 2.398 | 0.025 | 0.126 |
| I feel pessimistic. | 1.91 | 1.57 | 1.624 | 0.119 | 0.227 |
| I find it hard to see the value in living. | 1.65 | 1.35 | 1.576 | 0.129 | 0.236 |
| I feel like a loser. | 1.52 | 1.22 | 2.612 | 0.016* | 0.126 |
| I feel angry. | 1.22 | 1.17 | 0.439 | 0.665 | 0.698 |
| I feel like I'm boiling inside. | 1.09 | 1.04 | 1 | 0.328 | 0.417 |
| I feel indignant. | 1.09 | 1.04 | 1 | 0.328 | 0.417 |
| I feel so angry I could lose control. | 1.09 | 1.04 | 1 | 0.328 | 0.417 |
| I feel irritable. | 1.35 | 1.13 | 2.011 | 0.057 | 0.171 |
| I feel uncomfortable. | 1.3 | 1.43 | -0.617 | 0.544 | 0.601 |
| I feel outraged. | 1.09 | 1.04 | 1 | 0.328 | 0.417 |
| I act out in anger. | 1.74 | 1.65 | 0.385 | 0.704 | 0.721 |
| I easily get angry. | 2.04 | 1.57 | 1.751 | 0.094 | 0.208 |
| I behave aggressively. | 1.78 | 1.57 | 1.096 | 0.285 | 0.417 |
| I have trouble controlling my anger. | 1.78 | 1.48 | 1.432 | 0.166 | 0.279 |
| I tend to yell at others when I'm angry. | 1.83 | 1.48 | 1.624 | 0.119 | 0.227 |
| I flare up over trivial matters. | 2 | 1.57 | 2.647 | 0.015* | 0.126 |
| I say hurtful things when I'm angry. | 1.83 | 1.7 | 0.826 | 0.418 | 0.488 |
| I worry about trivial things. | 3 | 1.96 | 3.425 | 0.002* | 0.042* |
| I can't shake off bad thoughts when they come. | 2.61 | 1.96 | 2.343 | 0.029* | 0.126 |
| When I get anxious, I feel out of control. | 2.13 | 1.91 | 0.722 | 0.478 | 0.543 |
| I feel sensitive. | 3.17 | 2.52 | 1.875 | 0.074 | 0.173 |
| I feel impatient when I'm late for an appointment. | 3.74 | 2.91 | 2.455 | 0.022* | 0.126 |
| I get agitated easily. | 2.52 | 2.04 | 1.91 | 0.069 | 0.173 |
| I get distressed over minor things. | 2.74 | 2.26 | 1.665 | 0.11 | 0.227 |
| I feel gloomy. | 2.17 | 1.74 | 2.328 | 0.03* | 0.126 |
| I feel depressed. | 2 | 1.65 | 1.886 | 0.073 | 0.173 |
| I feel bothered by daily life. | 2.43 | 2.22 | 0.926 | 0.365 | 0.451 |
| I lack self-confidence. | 2.57 | 2.13 | 1.226 | 0.233 | 0.362 |
| Nothing interests me. | 2.04 | 1.74 | 1.321 | 0.2 | 0.323 |
| I feel lethargic. | 2.39 | 2.3 | 0.336 | 0.74 | 0.74 |
| The world feels dull and meaningless. | 1.96 | 1.83 | 0.53 | 0.601 | 0.647 |

### 3. Prefrontal Cortex Activation (fNIRS)

#### tcVNS condition vs Resting condition

To assess cortical hemodynamic changes induced by tcVNS, Wilcoxon signed-rank tests were conducted for each fNIRS channel, comparing oxygenated (HbO) and deoxygenated hemoglobin (HbR) signals between the tcVNS and resting conditions. Several channels exhibited significant differences in HbO concentration. In particular, Channel 08 (right frontopolar prefrontal cortex) showed a significant reduction in HbO during tcVNS (z = –2.44, p = .0146), while Channel 14 (right orbitofrontal cortex) showed a significant increase (z = 3.17, p = .0015). Additional significant changes were observed in Channel 23 (left frontopolar prefrontal cortex; z = –2.06, p = .0392) and Channel 42 (left frontopolar prefrontal cortex; z = 2.06, p = .0392), indicating regionally heterogeneous effects of stimulation.

In the HbR signal, a corresponding pattern was evident: Channel 08 (right frontopolar prefrontal cortex) showed a significant increase in HbR (z = 2.50, p = .0125), while Channel 14 (right orbitofrontal cortex) displayed a significant decrease (z = –3.23, p = .0012), suggesting reduced oxygen consumption or increased local clearance. Channels 23 and 42 (both in the left frontopolar prefrontal cortex) also showed statistically significant HbR changes (z = 2.20, p = .0277 and z = –1.99, p = .0464, respectively). These findings suggest that tcVNS modulates cortical blood oxygenation, particularly in the bilateral frontopolar and right orbitofrontal cortices, which may be functionally relevant to autonomic or affective regulation. Future work should map these channels onto anatomical landmarks for more precise interpretation (see Figure 11).

**Figure 11.**
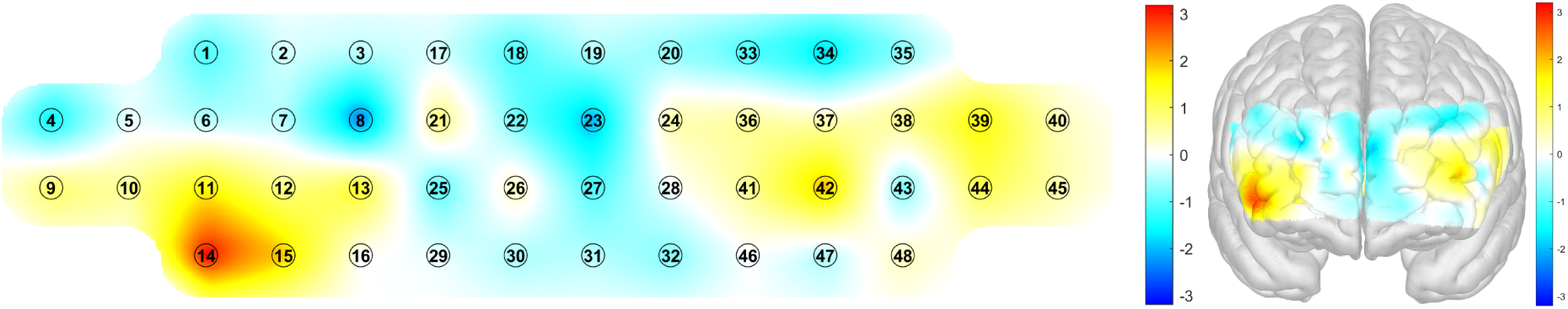
Cortical hemodynamic changes during tcVNS relative to resting: fNIRS channel and brain surface activation maps.

#### tcVNS+Breathing+Aroma condition vs. Breathing+Aroma condition

To evaluate the additional effect of tcVNS when combined with breathing and aroma stimulation, Wilcoxon signed-rank tests were conducted for each fNIRS channel comparing hemodynamic responses between the combined multimodal condition and the Breathing + Aroma condition. Wilcoxon signed-rank tests were conducted to compare cortical hemodynamic activity between the tcVNS+Breathing+Aroma condition and the Breathing+Aroma condition. Analysis of oxygenated hemoglobin (HbO) revealed that Channel 01 (right dorsolateral prefrontal cortex) showed a statistically significant reduction in HbO during the tcVNS-integrated condition (z = –2.20, p = .028). Channel 31, corresponding to the left orbitofrontal cortex (lOFC), also demonstrated a significant increase (z = 2.04, p = .0409). Marginally significant increases were observed in additional regions: Channel 20 (left DLPFC, z = 1.96, p = .0505), Channel 28 (left frontopolar prefrontal cortex, z = 1.78, p = .0747), and Channel 03 (right DLPFC, z = 1.76, p = .0796).

A significant reduction in HbR was observed in Channel 31, corresponding to the left orbitofrontal cortex (z = –2.13, p = .033), suggesting increased local activation under the combined stimulation. Similarly, Channel 01 (right dorsolateral prefrontal cortex) showed a significant increase in HbR (z = 2.20, p = .028), possibly indicating regional suppression or compensatory redistribution. Marginally significant effects were also found in Channel 20 (left dorsolateral prefrontal cortex, z = –1.78, p = .075), Channel 03 (right dorsolateral prefrontal cortex, z = –1.75, p = .080), and Channel 28 (left frontopolar prefrontal cortex, z = –1.71, p = .087), reflecting potential trends toward HbR modulation with combined tcVNS involvement.

While none of these effects survived FDR correction, the pattern of HbR changes suggests spatially distributed hemodynamic sensitivity to vagal stimulation when combined with slow breathing and olfactory input, particularly in lateral and medial prefrontal areas involved in affective and autonomic integration. Taken together, these findings suggest that tcVNS, both alone and in combination with other interventions, engages distinct prefrontal circuits involved in emotional, cognitive, and autonomic regulation, with the combined condition potentially reflecting the recruitment of top-down cognitive control and autonomic regulation networks (see Figure 12).

**Figure 12.**
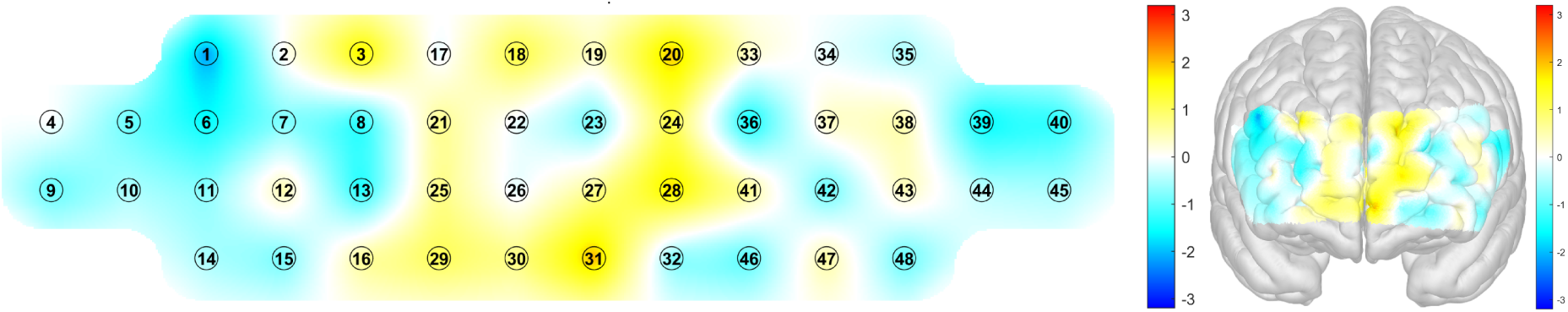
Cortical hemodynamic changes during tcVNS relative to resting: fNIRS channel and brain surface activation maps.

## Discussion

This study demonstrated that transcutaneous cervical vagus nerve stimulation (tcVNS), slow breathing with aromatherapy, and especially their combination each produced significant autonomic and emotional benefits compared to the resting baseline. All active conditions yielded substantial increases in HRV time-domain metrics, with SDNN showing robust gains under tcVNS and even greater enhancement when combined. RMSSD followed a similar trend, with the combined intervention outperforming either modality alone. These parasympathetic improvements coincided with marked reductions in self-reported anxiety and worry over the course of the session. In parallel, functional near-infrared spectroscopy (fNIRS) revealed that both tcVNS alone and its combination with breathing and aroma elicited significant changes in prefrontal cortical oxygenation. Specifically, tcVNS alone increased oxyhemoglobin in right orbitofrontal cortex while decreasing it in bilateral frontopolar areas, whereas the combined condition (vs breathing+aroma) produced elevated oxyhemoglobin in left orbitofrontal cortex and reduced oxyhemoglobin in right dorsolateral PFC. These cortical effects suggest engagement of top-down regulatory networks. Taken together, the pattern of results – greater HRV and complexity, lower stress, and modulated prefrontal activity – is consistent with the restoration of vagal tone and enhanced prefrontal regulation. In line with the neurovisceral integration model, improved HRV likely reflects strengthened parasympathetic control over cardiac function, concomitant with increased prefrontal influence over subcortical stress circuits (Thayer & Friedman, 2004). Importantly, the combined tcVNS+breathing+aroma protocol produced the most robust improvements in both physiological and emotional measures. This synergy implies that convergent multimodal inputs (vagal, respiratory, olfactory) yield additive benefits beyond any single intervention, fulfilling our hypothesis that a triple-modality approach would maximize autonomic balance and stress relief.

### Autonomic Effects of tcVNS

Transcutaneous vagus nerve stimulation alone produced clear signs of enhanced parasympathetic activity. Time-domain HRV indices improved markedly: SDNN showed robust gains with tcVNS, and RMSSD also rose substantially, nearing statistical significance. The proportion of successive NN intervals differing by more than 50 ms increased, indicating greater high-frequency variability. Non-linear measures shifted in a vagal direction as well: SD1 increased, and approximate entropy (ApEn) climbed, suggesting that heart rate patterns became more complex and adaptable under tcVNS, reflecting a more flexible autonomic system. In summary, tcVNS reliably elevated HRV across multiple metrics, signifying a shift toward vagal dominance. These autonomic changes align with established vagal mechanisms. Electrically stimulating the auricular vagus branch activates afferent fibers projecting to the nucleus tractus solitarius, ultimately enhancing efferent vagal outflow and inhibiting sympathetic tone (Deuchars et al., 2018). The observed increases in HRV echo findings from previous tVNS studies: for example, Höper et al. (2022) reported significant HRV augmentation (and heart rate reduction) under tVNS, coupled with concurrent increases in prefrontal oxygenation (Höper, Kaess, & Koenig, 2022). In our data, the tcVNS-evoked HRV enhancements mirror those reports, reinforcing the view that vagal afferent activation can quickly improve cardiac autonomic regulation. Notably, these changes emerged in healthy participants during brief stimulation, indicating that taVNS can acutely elevated vagal tone even in the absence of pathology (Forte et al., 2022).

The increase in HRV indices under tcVNS suggests improved baroreflex function as well. Although we did not directly measure baroreflex sensitivity, prior work has shown that noninvasive VNS enhances baroreceptor function (Antonino et al., 2017) and that slow exhalatory breathing gates vagal impulses to boost cardiovagal tone (Laborde et al., 2021). The trend toward lower heart rate with tcVNS (as seen in related studies) would be consistent with enhanced baroreflex-mediated parasympathetic activity (Antonino et al., 2017). Moreover, our finding of elevated HRV complexity (ApEn) implies that the cardiac control system became more responsive and less rigid (Sarkar & Barat, 2006). In the context of neurovisceral integration, higher HRV reflects a more adaptive central-autonomic network (Thayer & Lane, 2000). Thus, the autonomic data provide strong evidence that tcVNS alone can restore vagal modulation of the heart, setting the stage for improved emotion regulation and stress resilience via central pathways.

### Multimodal Synergy

The combined intervention of tcVNS with paced breathing and aromatherapy produced synergistic enhancements that surpassed any single treatment. Most strikingly, heart rate variability under the combined protocol was substantially greater than with either tcVNS alone or breathing plus aroma. Likewise, RMSSD showed its largest increase when all three modalities were applied together, exceeding the improvements seen with each modality by itself. Statistical analyses confirmed that these combined effects were highly significant. Subjective stress ratings mirrored this pattern: participants reported the reduction in stress following the triple treatment. Altogether, these results demonstrate that integrating electrical stimulation, controlled breathing, and olfactory cues yields additive—and in some cases more-than-additive—benefits for autonomic regulation and mood. This synergy likely arises from complementary bottom-up and top-down mechanisms. Slow, 6-breaths/min breathing is known to engage the baroreflex at its resonant frequency, greatly amplifying cardiac vagal efference (Larson et al., 2020). It also recruits cortical attention networks when one focuses on the breath. Aromatic lavender/bergamot inhalation provides a sensory-affective stimulus that directly activates limbic regions (olfactory bulb, amygdala) and orbitofrontal networks (Watanabe et al., 2015). Combining these with tcVNS means that at each respiratory cycle, one is simultaneously sending strong afferent vagal signals (via VNS) and attenuated baroreceptor bursts (via exhalation). Conceptually, this is similar to “respiratory-gated VNS” (RAVANS) approaches, where timing vagus pulses to exhalation has been shown to enhance cardiovagal tone (Sclocco et al., 2017). Although our study used continuous VNS, the overlap of signals would be maximal during exhalation. Thus, breathing and aroma may prime or boost the brainstem circuits that VNS targets, leading to greater vagal outflow. In short, slow paced breathing increases the substrate on which VNS acts, and pleasant odors modulate affective circuitry to further bias the system toward relaxation. The result is a convergence of multiple autonomic inputs (vagal, baroreflex, olfactory) and attentional focus, creating a “force-multiplier” effect on parasympathetic activation and stress reduction.

These findings extend previous work by showing the benefits of simultaneous multisensory intervention. Prior studies have reported improvements from individual components — for example, aromatherapy alone can raise high-frequency HRV and lower cortisol (Chien et al., 2012), and tVNS alone can lower sympathetic arousal in stress contexts (Gurel et al., 2020). However, reports of additive effects are scarce. Our data suggest that when combined, the interventions interact constructively. From a neurophysiological standpoint, this makes sense: the nucleus tractus solitarius and related brainstem nuclei receive convergent inputs from vagal afferents, baroreceptors, and higher centers (Bonham & Hasser, 1993). Furthermore, higher-level networks may be more fully engaged: for instance, the prefrontal cortex may integrate interoceptive (breath, VNS) and exteroceptive (olfactory) cues, yielding a reinforced sense of safety (Lefranc et al., 2020). In summary, the triple condition’s superior outcomes imply true multimodal synergy, consistent with a polyvagal/neurovisceral perspective in which converging neural inputs stabilize affective and autonomic states.

### Cortical Effects (PFC)

Functional imaging revealed that tcVNS and the combined protocol modulated prefrontal cortical activity. During tcVNS alone, we observed significant changes in the frontopolar and orbitofrontal cortices. In particular, tcVNS significantly increased oxyhemoglobin in the right orbitofrontal cortex while decreasing oxyhemoglobin in bilateral frontopolar regions. When comparing the combined protocol to breathing plus aroma alone, the most pronounced effects appeared in dorsolateral and orbitofrontal areas, with the combined treatment boosting oxygen levels in the left orbitofrontal cortex while reducing them in the right dorsolateral prefrontal cortex. Altogether, these shifts indicate that the interventions recruited a network of prefrontal regions involved in both executive function and emotional regulation. The increased oxygenation in the orbitofrontal cortex likely reflects active emotion regulation. The OFC is implicated in evaluating affective value and modulating amygdala responses (Fettes, Schulze, & Downar, 2017; Thayer et al., 2012), so higher OFC engagement during tcVNS and the combined condition suggests greater top-down inhibition of limbic arousal. Similarly, frontopolar and dorsolateral PFC changes may relate to altered cognitive control demands. The observed decreases in some frontal areas (e.g. right DLPFC under the combined intervention) could indicate reduced cortical effort as autonomic balance was restored, or alternatively reflect an inhibitory process. Importantly, all these frontal cortical changes occurred in the same directions expected from the neurovisceral integration framework: enhanced parasympathetic tone (higher HRV) is typically accompanied by increased functional connectivity and modulation in medial and orbital PFC regions (Thayer & Lane, 2000). For instance, Hӧper et al. found that under tVNS, relative increases in HRV were correlated with rising oxygenation in lateral and medial PFC over time (Höper et al., 2022). Our results are consistent with that, indicating that vagal stimulation promotes prefrontal recruitment.

Overall, the PFC effects support the idea that these interventions operate via cortical autonomic regulatory circuits. The orbitofrontal and frontopolar cortices are part of the limbic-associative network that exerts inhibitory control over subcortical stress centers (Fettes et al., 2017). By modulating blood flow in these areas, tcVNS and the combined treatment likely tap into a cortical gate on emotion and autonomic output. This is in keeping with the neurovisceral model: the PFC (particularly medial and orbital regions) modulates the amygdala and brainstem via vagal pathways (Thayer & Lane, 2000). Thus, our fNIRS findings provide a cortical correlation for the observed HRV improvements and stress reductions, suggesting that the interventions strengthened prefrontal regulatory influence as autonomic homeostasis was restored (Sakaki, Yoo, & Mather, 2016).

### Subjective Stress & Emotion

Subjects reported clear mood improvements across the active interventions. Anxiety scores decreased significantly, and worry scores also fell. Participants also noted feeling less restlessness, gloominess, and unhappiness after the session. These subjective gains parallel the physiological changes: increasing evidence links enhanced vagal tone (higher HRV) with reduced anxiety and better emotional regulation (Goessl et al., 2017; Kim et al., 2018). In our data, the greatest HRV increases occurred with concomitant reductions in anxiety, consistent with this body–mind coupling. For example, the combined condition not only achieved the highest SDNN and RMSSD, but it was also under this condition that mood seemed most improved, suggesting that stronger parasympathetic activation translated into more relief from stress.

Aromatherapy likely contributed specifically to emotional relief. Essential oils like lavender and bergamot have well-documented anxiolytic and mood-lifting effects; they directly stimulate olfactory-limbic pathways to reduce stress hormones and boost parasympathetic indices (Watanabe et al., 2015). In our study, the breathing+aroma and combined sessions (which included scent) were associated with elevated HF-HRV (reflecting vagal influence) and with subjective comments of calmness. The Watanabe et al. (2015) trial, for example, found that inhaling bergamot oil significantly lowered salivary cortisol and increased HF-HRV in minutes, paralleling our pattern of immediate mood benefit. While we did not isolate aroma from breathing here, the general improvement across affective items suggests that sensory cues aided the relaxation response. Notably, the linkage between subjective and physiological data underscores the importance of mind–body integration (Thayer & Lane, 2000). The neurovisceral model predicts that higher PFC-mediated control (and thus HRV) coincides with better attentional and emotional regulation (Thayer et al., 2009). Our findings accord with this: individuals who achieved higher HRV also endorsed feeling calmer and more secure (Kim et al., 2018). This mutual reinforcement may create a positive feedback loop, whereby breathing focus and pleasant smells reduce anxiety, which in turn facilitates deeper vagal activation (Zaccaro et al., 2018). Taken together, the reduction in self-reported stress across conditions reflects the combined effects of physiological down-regulation and cognitive-affective shifts induced by the interventions.

### Comparative Literature & Mechanistic Interpretation

Our results extend and integrate insights from multiple lines of prior research. Transcutaneous vagus stimulation alone has been shown to dampen sympathetic responses in stress conditions (e.g. in PTSD patients) (Gurel et al., 2020) and to elevate HRV. Paced breathing at resonance frequency (∼6 breaths/min) is widely documented to spike HRV and baroreflex sensitivity (Lehrer & Gevirtz, 2014). Aromatherapy research has likewise reported acute increases in parasympathetic activity and mood improvement (Okada & Shimatani, 2024). However, most past studies examined these modalities in isolation. For instance, RAVANS (respiratory-gated VNS) studies have synchronized exhalation with auricular stimulation to target stress, showing moderate autonomic gains, but typically without added sensory stimuli. In contrast, our triple-combination approach is novel. To our knowledge, no previous investigation has combined tcVNS, slow breathing, and olfactory stimulation simultaneously to modulate stress.

Furthermore, our inclusion of cortical imaging is distinctive. Few tVNS studies have directly measured prefrontal oxygenation during stimulation; Hӧper et al. (2022) is an exception, showing that tVNS increases PFC oxygenation in adolescents. Here, by adding breathing and aroma, we were able to observe amplified PFC effects. This multi-sensory integration approach also builds on complementary literature in complementary medicine. For example, dual-modality relaxation interventions (e.g., music + breathing, or visuals + aroma) have been reported to yield greater effects than either alone, although these have rarely been quantified with HRV and fNIRS (Al-Shargie et al., 2022). Our findings thus highlight the value of a holistic, multi-input strategy. In sum, compared with prior single-domain studies, the present work offers a more powerful intervention paradigm and provides evidence that simultaneous bottom-up (vagal, baroreceptor, olfactory) and top-down (attentional) inputs can jointly enhance stress regulation.

The physiological and cortical effects observed can be integrated into a coherent mechanistic framework. TcVNS delivers electrical stimulation to afferent vagal fibers in the neck, which project to the nucleus tractus solitarius (NTS) in the brainstem. From the NTS, signals are relayed to the dorsal motor nucleus of the vagus and nucleus ambiguus to increase parasympathetic output to the heart (Fang et al., 2023). The NTS also projects to neuromodulatory centers such as the locus coeruleus and forebrain regions including the prefrontal cortex (Thayer & Lane, 2000), thereby influencing broader regulatory networks. This pathway explains how tcVNS lowers heart rate and elevates HRV.

Meanwhile, paced exhalation at ∼6 breaths/min engages arterial baroreceptors on each exhale, sending rhythmic bursts to the NTS that further amplify vagal efference. Slow breathing thus acts through a baroreflex mechanism to magnify heart–brain coupling (Larson et al., 2020). The convergence of these pathways at the NTS may produce a summative effect: our combined condition effectively delivered continuous vagal stimulation (tcVNS) together with phasic baroreflex inputs (breathing), so the brainstem autonomic network received strongly concordant “relaxation” signals from both sources.

Concurrently, aromatherapy influences the system via olfactory pathways. Pleasant scents like lavender and bergamot are detected by olfactory receptors, whose signals travel to the olfactory bulb and then to limbic structures such as the amygdala, hippocampus, and orbitofrontal cortex (Herz, 2009). Activation of these areas can modulate the hypothalamus and brainstem centers to reduce stress hormone release and sympathetic tone. In other words, aroma provides a direct limbic afferent that biases the system toward calm by attenuating amygdala-driven arousal (Hossein et al., 2005). Notably, the orbitofrontal cortex — which showed increased activation in our fNIRS data — is a primary cortical target of olfactory input, suggesting that scent may have amplified cortical regulatory feedback.

At the cortical and interoceptive level, the prefrontal cortex and insula likely integrate these bottom-up signals (Critchley & Harrison, 2013). The vmPFC/OFC can inhibit the amygdala and modulate autonomic responses; increased OFC oxygenation (as seen under tcVNS and the combined condition) implies engagement of this top-down brake (Beissner et al., 2013). The insular cortex, representing body states, would register the changes in heart rate and breathing, and provide feedback to PFC about the internal milieu. Thus, a PFC–insula–amygdala network may be dynamically rebalanced by the interventions: enhanced vagal tone and pleasant olfactory input suppress amygdala fear activity, while PFC/insula detect and reinforce the signal that “all is well,” sustaining parasympathetic dominance. In sum, tcVNS, breathing, and aroma appear to co-activate interrelated neurocircuits (NTS–vagal pathways, baroreflex loops, olfactory–limbic pathways, and prefrontal control circuits) that collectively stabilize heart–brain function. The robust HRV increases and cortical shifts we observed reflect this multi-level integration of autonomic and affective regulation.

### Clinical Translation & Future Directions

The present findings have promising implications for clinical stress management. Non-invasive tcVNS and behavioral techniques like paced breathing and aromatherapy could be combined into a portable or desktop intervention for anxiety, PTSD, depression, or cardiovascular patients, aiming to restore autonomic balance (Badran et al., 2019). Elevated HRV and lowered anxiety in our healthy sample suggest that similar protocols might benefit individuals with autonomic dysregulation or affective disorders (Zaccaro et al., 2018). For example, patients with PTSD exhibit chronically low HRV and heightened sympathetic tone; targeted tcVNS protocols have shown preliminary success in such populations (Gurel et al., 2020). A wearable VNS device paired with guided breathing exercises and calming scents could potentially be used adjunctively in therapy or mindfulness training to enhance resilience.

However, translation to practice requires further research.Future studies should employ rigorous sham-controlled, randomized designs to isolate the specific effects of each component and control for expectancy. The optimal parameters (stimulation frequency, intensity, breathing rate, duration) need to be established. Longitudinal trials are needed to determine whether repeated sessions produce cumulative benefits or longer-lasting changes in trait anxiety and cardiovascular health (Yap et al., 2020). It would also be valuable to test efficacy in diverse groups: for instance, adolescents (who have developing autonomic control) and older adults (who often have reduced HRV) could respond differently (Koenig et al., 2016). Moreover, exploring personalization (e.g. adjusting breathing rate to each individual’s resonance frequency) might maximize effectiveness. Future work could also incorporate real-time feedback: measuring HRV live to titrate stimulation or breathing rate could create a closed-loop biofeedback system (Shaffer & Meehan, 2020).

Finally, our use of fNIRS to monitor PFC engagement opens avenues for neurofeedback: individuals might learn to modulate frontal activation alongside breathing and VNS. In summary, while the current single-session results are encouraging, robust clinical trials with active and sham comparisons, varied populations, and optimized protocols are necessary before routine application. If validated, multimodal interventions like this could offer a drug-free, side-effect-free strategy for mental health and stress-related conditions, leveraging the brain–heart connection for therapeutic gain.

## Data Availability

All data produced in the present study are available upon reasonable request to the authors

## Acknowledgements

This work was supported by the Yonsei Signature Research Cluster Program (Project title: “Psychological Prevention for the Untact University Generation: Longitudinal Psychological Health Database and Multidimensional Psycho-Social Functioning Prediction Model”).

